# Electroencephalographic Transient Beta Event Rates in Autism and Related Neurogenetic Conditions

**DOI:** 10.64898/2026.08.02.26359366

**Authors:** Gerardo Parra, Klara Szilagyi, Yael Braverman, Devorah Kranz, E. Martina Bebin, Raphael A. Bernier, Jonathan A. Bernstein, Elizabeth Berry-Kravis, Joseph D. Buxbaum, Katarzyna Chawarska, Emma E. Condy, Laura Cornelissen, Geraldine Dawson, Abigail Dickinson, James Dziura, Charis Eng, Susan Faja, Jennifer H. Foss-Feig, Jeffrey P. Gavornik, Antonio Y. Hardan, Ellen Hanson, Shafali Jeste, Linnea Joffe-Nelson, Alexander Kolevzon, Darcy A. Kreuger, Balu Krishnan, Robert Law, David N. Lieberman, Claire MacKay, Julian A. Martinez-Agosto, Adam J. Naples, Charles A. Nelson, Hope Northrup, Ernest Pedapati, Annapurna Poduri, Vineet Punia, Rajsekar R. Rajaraman, David M. Ritter, Celine Saulnier, Frederick Shic, Latha Valluripalli Soorya, Catherine A. Sugar, Kyle E. Takach, Audrey Thurm, Sara J. Webb, Kimberly Wiltrout, Damla Şentürk, Stephanie Jones, Michela Fagiolini, James C. McPartland, Mustafa Sahin, April R. Levin, Developmental Synaptopathies Consortium, Autism Biomarkers Consortium for Clinical Trials

**Affiliations:** Department of Neurology, Boston Children’s Hospital/Harvard Medical School, Boston, MA, USA; Rosamund Stone Zander and Hansjörg Wyss Translational Neuroscience Center, Boston Children’s Hospital, Boston, MA, USA; Program in Neuroscience, Division of Medical Sciences, Graduate School of Arts and Sciences, Harvard Medical School, Boston, MA, USA; University of Alabama School of Medicine, Birmingham, AB, USA; Department of Psychiatry and Behavioral Sciences, University of Washington, Seattle, WA, USA; Department of Pediatrics, Stanford University School of Medicine, Stanford, CA, USA; Rush University Medical Center, Chicago, IL, USA; Seaver Autism Center for Research and Treatment, Icahn School of Medicine at Mount Sinai, New York, NY, USA; Department of Psychiatry, Icahn School of Medicine at Mount Sinai, New York, NY, USA; Department of Genetics and Genomic Sciences, Icahn School of Medicine at Mount Sinai, New York, NY, USA; Department of Neuroscience, Icahn School of Medicine at Mount Sinai, New York, NY, USA; Child Study Center, School of Medicine, Yale University, New Haven, CT, USA; Neurodevelopmental and Behavioral Phenotyping Service, National Institutes of Health, Bethesda, MD, USA; Department of Anesthesiology, Critical Care & Pain Medicine, Boston Children’s Hospital/Harvard Medical School, Boston, MA, USA; Current Address: Eisai Inc., Clinical Evidence Generation – Neurology, Nutley, NJ, USA; Duke Institute for Brain Sciences, Duke University, Durham, NC, USA; Duke Center for Autism and Brain Development, Duke University, Durham, NC, USA; Department of Psychiatry and Behavioral Sciences, Duke University, Durham, NC, USA; Semel Institute of Neuroscience and Human Behavior, David Geffen School of Medicine, University of California, Los Angeles, CA, USA; Yale Center for Clinical Investigation, Yale University, New Haven, CT, USA; Yale Center for Analytical Sciences, Yale University, New Haven, CT, USA; Genomic Medicine Institute, Lerner Research Institute, Cleveland Clinic, Cleveland, OH, USA; Center for Personalized Genetic Healthcare, Medical Specialties Institute, Cleveland Clinic, Cleveland, OH, USA; Department of Genetics and Genome Sciences, Case Western Reserve University School of Medicine, Cleveland, OH, USA; Department of Molecular Medicine, Cleveland Clinic Lerner College of Medicine of Case Western Reserve University, Cleveland, OH, USA; Division of Developmental Medicine, Boston Children’s Hospital, Harvard Medical School, Boston, MA, USA; Department of Psychiatry and Behavioral Sciences, Stanford University, Stanford, CA, USA; Department of Neurology, University of Southern California Keck School of Medicine, Los Angeles, CA, USA; Department of Pediatrics, University of Southern California Keck School of Medicine, Los Angeles, CA, USA; Department of Pediatrics, Icahn School of Medicine at Mount Sinai, New York, NY, USA; Division of Neurology, Cincinnati Children’s Hospital Medical Center, Cincinnati, OH, USA; Department of Pediatrics, University of Cincinnati College of Medicine, Cincinnati, OH, USA; Neurological Institute, Epilepsy Center, Cleveland Clinic, Cleveland, OH, USA; Department of Human Genetics, David Geffen School of Medicine, University of California, Los Angeles, CA, USA; Department of Pediatrics, David Geffen School of Medicine, University of California, Los Angeles, CA, USA; Department of Psychiatry, David Geffen School of Medicine, University of California, Los Angeles, CA, USA; Department of Human Genetics and Department of Pediatrics, University of California, Los Angeles, CA, USA; Department of Pediatrics, McGovern Medical School, University of Texas Health Science Center at Houston (UTHealth) and Children’s Memorial Hermann Hospital, Houston, TX, USA; Department of Psychiatry, Cincinnati Children’s Hospital Medical Center, University of Cincinnati College of Medicine, Cincinnati, OH, USA; Epilepsy Center, Cleveland Clinic, Cleveland, OH, USA; Division of Pediatric Neurology, Department of Pediatrics, UCLA Mattel Children’s Hospital, David Geffen School of Medicine, Los Angeles, CA, USA; Neurodevelopmental Assessment and Consulting Services, Decatur, GA, USA; Center for Child Health, Behavior and Development, Seattle Children’s Research Institute, Seattle, WA, USA; Department of Pediatrics, University of Washington, Seattle, WA, USA; Department of Biostatistics, University of California, Los Angeles, CA, USA; Department of Psychiatry and Biobehavioral Sciences, University of California, Los Angeles, CA, USA; Stritch School of Medicine, Loyola Medical School, Maywood, IL, USA; Department of Neuroscience, Brown University, Providence, RI, USA; Center for Neurorestoration and Neurotechnology, Providence VA Medical Center, Providence, RI, USA; Hock E. Tan and K. Lisa Yang Center for Autism Research at Harvard University, Boston, MA, USA; International Research Center for Neurointelligence (IRCN), University of Tokyo Institutes for Advanced Study, Tokyo, Japan; Yale Center for Brain and Mind Health, Yale University, New Haven, CT, USA

**Author notes:** These authors contributed equally to this work (Gerardo Parra, Klara Szilagyi, Yael Braverman).

## Abstract

Transient beta events (TBE) during electroencephalography (EEG) reflect thalamocortical activity, bridging genotype to phenotype and impacting sensory responsivity. Compared to typically developing controls, we found elevated TBE rate in some children with idiopathic Autism Spectrum Disorder (ASD) and a majority of children with Phelan-McDermid Syndrome, Rett Syndrome, and *SYNGAP1*-related disorder. TBE rate thus offers promise as a stratification biomarker with divergent and convergent properties across ASD and neurogenetic conditions, respectively.

## Main Text

Autism spectrum disorder (ASD) is a prevalent and heterogeneous neurodevelopmental condition.^1^ Translational biomarkers in ASD and other neurodevelopmental disorders are crucial for clinical trial readiness and reflect efforts to understand the circuit-level neurobiology bridging genotype to phenotype^2^. Electroencephalography (EEG) offers particular promise in this regard, given its high temporal resolution and scalability^3^. While biomarkers can be considered for multiple contexts of use in ASD, stratification is a particular priority because of the vast heterogeneity inherent to ASD. Identifying potential EEG-based biomarkers that converge across some genetically-defined disorders in which ASD is common, but are present only in a subset of children with the ASD phenotype, offer opportunities to inform meaningful stratification based on shared mechanisms of neural circuit function (i.e., a bridge between genotype and phenotype)^4^. Biomarkers for which the underlying mechanisms are well-described, and those that are related to a meaningful clinical outcome, offer particular benefit in supporting clinical trial readiness and translation of targeted treatments from bench to bedside.

Atypical sensory responsivity ^5, 6^ is common in autistic individuals and animal models of ASD ^7–14^. In humans, sensory differences can impede daily activities and negatively affect quality of life, leading to downstream increases in anxiety, depression, repetitive behaviors, and attention problems ^10, 15^. Improving sensory processing challenges is repeatedly noted as a common goal that would improve quality of life^16, 17^. Current assessment of sensory responsivity traditionally relies on subjective self, parent, or clinician report, and views hyper- and hypo-responsivity as opposing traits. However, the “Sensory Paradox” recently described that hyper- and hypo-responsivity tend to co-occur within individuals and are positively correlated with one another^18^.

Transient Beta Events (TBEs) offer particular potential as a mechanistically relevant biomarker^19^ that may help to explain the Sensory Paradox in ASD. TBEs are spontaneous, transient (typically <150ms) increases in high power cortical activity in the beta frequency band (13-30 Hz), that are visible in resting EEG (i.e., EEG in the awake resting state with no time-locked stimuli), magnetoencephalography (MEG), and local field potential (LFP) recordings ^20, 21^. Cross-species studies and computational models suggest that TBEs reflect underlying thalamocortical circuit activity, arising from a broad burst of excitatory synaptic drive from lemniscal thalamus to the proximal dendrites of cortical pyramidal neurons in the infragranular layers that occurs simultaneously with a brief (∼50 ms), stronger burst of drive from nonlemniscal thalamus to distal pyramidal neuron dendrites in the supragranular layers. ^21, 22^. TBEs are found in many cortical regions, have been studied in animal models and neurotypical humans ^20–24^ and are associated with behavioral measures of sensory perception ^21, 24^, as well as memory ^25, 26^ and movement-related tasks ^23, 27–30^. While traditional measurements of beta represent frequency bounded average spectral power, TBEs are quantified according to how often they occur and “TBE rate” represents event frequency in Hz. Unlike more traditional metrics of averaged beta power, TBEs are quantified using unaveraged data and provide finer grain insights into the functional relationship between spontaneous, intrinsic neural processes and discrete sensory and behavioral events ^26, 27^.

Tactile detection experiments in the somatosensory cortex provide an example of the potential for granular TBE analysis to link EEG with underlying neural processes and help explain the “Sensory Paradox.”^18^. When near-detection threshold stimuli are delivered to a finger, behaviorally assayed detection probability can increase or decrease across trials within an individual, depending on the exact timing relationship between the tactile stimulus and the TBE. Specifically, detection probability increases on a trial-by-trial basis when a TBE overlaps with tactile stimulation (e.g. stimulation precedes the TBE peak by up to ∼ 50 ms) and decreases when TBEs precede stimulation by up to ∼200 ms ^21, 22, 24^. These results, bolstered by similar reports in other brain regions ^31–33^, demonstrate that TBEs serve a complex gating function at the intersection of sensory processing and high-order cognitive functions thought to be involved with phenotypic trait expression in ASD ^34^. The bidirectional modulation of perceptual detection and accessibility of EEG make TBEs particularly intriguing for ASD research, where differences in tactile processing express as both hyper- and hypo-responsivity ^12^. Despite compelling evidence that TBE analysis provides a lens into neurophysiological mechanisms underlying sensory responsivity and cascading cognitive and behavioral effects thereof^15^, no study has yet characterized TBE rates in ASD or related neurodevelopmental conditions.

The primary goals of this study were therefore to use resting EEG (1) to determine whether TBE rates differ between typically developing (TD) and ASD participants; (2) to understand whether TBE rates could align with genetically defined subtypes by evaluating TBE rates in 5 neurogenetic conditions associated with ASD and related neurodevelopmental conditions; (3) to evaluate the short-term reliability of TBE rates to begin considering their potential clinical utility; and (4) to explore the relationship between spontaneous TBE rates and tactile responsivity.

Demographics and clinical characteristics of transdiagnostic, multi-site participants are reported in Tables S1 and S2; see Methods section for study details. TD participants were recruited from these studies and studies of 5 neurogenetic conditions described below (n=180; Table S1).

Matching the previous focus on tactile processing^22, 24^, primary analyses focused on TBE rates (Hz) from electrodes over somatosensory cortex (C3/C4; Table S3). For completeness, full topoplots are included in Figure S1. Since TBE rates did not differ by study for TD participants (Figure S2), we collapsed them into one group.

For goal (1), we evaluated resting EEGs recorded in two studies of idiopathic ASD: the “Sensory Processing and Adaptation” (SPA) study of sensory processing in preschoolers (ASD_SPA; n=26), and the “Autism Biomarkers Consortium for Clinical Trials” ^35^ study of social communication biomarkers in school-aged children (ASD_ABC-CT; n=262). We tested whether TBE rates in ASD participants from ASD_SPA and/or ASD_ABC-CT differed from TD (Figure 1a). TBE rates were higher in ASD_SPA (U =3867, p<.001, r=0.29) but not ASD_ABC-CT relative to TD (U=26461, p=.127, r=0.07).

**Figure 1.**
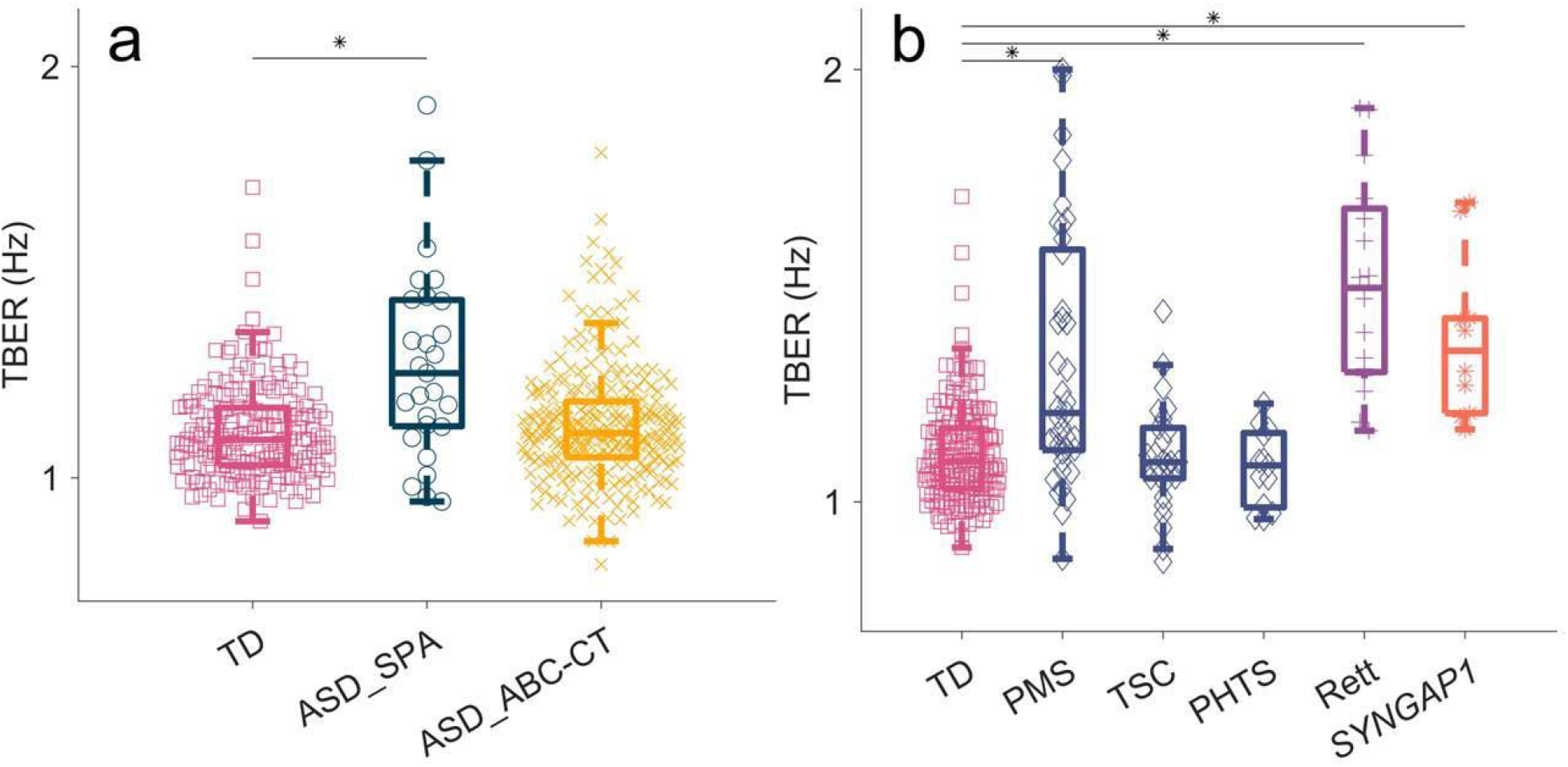
Transient Beta Event Rate measured over electrodes C3/C4 across diagnostic groups. Each shape reflects one participant. Within each box, horizontal middle line denotes group median; boxes extend over each group’s interquartile-range (IQR); vertical dashed whiskers denote values 1.5 times the IQR value. (a) * indicates significant group differences assessed using a Mann-Whitney U test (p < .001). TBE rates did not differ between TD and ASD_ABC-CT (p = .254). (b) * indicates significant group differences assessed using repeated Mann-Whitney U tests (p < .001, Bonferonni corrected for five comparisons). TBE rates did not significantly differ between TD and TSC or PHTS (both adjusted p = 1.0)

For goal (2), considering the heterogeneity of ASD and the findings above, we tested whether TBE rates could differentiate sub-groups in five neurogenetic conditions in which ASD is common: Phelan McDermid Syndrome (PMS; n=41), Tuberous Sclerosis Complex (TSC; n=25), PTEN Hamartoma Tumor Syndrome (PHTS; n=12), Rett syndrome (RTT; n=15), and SYNGAP1-Related Disorder (SYNGAP1; n=11). TBE rate was elevated (Figure 1b, all p <.001) in PMS (U=5547.5, r=0.32), RTT (U=3630, r=0.49), and SYNGAP1 (U=1645, r=0.32) participants relative to TD but not in TSC (U=2280, p=.833, r=0.01) or PHTS (U=1000.5, p=.383, r=0.06). TBE rates did not differ based on co-occurring ASD diagnostic status (Table S2; U=883.5, p = .379). These results show that TBE rates are higher in a subset of ASD-related neurogenetic conditions (Figure 1), but a clinical diagnosis of ASD itself cannot account for elevated TBE rates.

For goal (3), to begin considering potential clinical utility of TBE rates, we next evaluated short-term reliability using data from the ABC-CT cohort collected six weeks post initial visit. TBE rates short-term reliability was poor for the TD group, (ICC .365, 95% CI [.189, .517]) but good for the ASD group (ICC .606, 95% CI [.520, .679]) (Figure S3).

For goal (4), we used the Sensory Profile 2 (SP2^23^), a caregiver-reported questionnaire completed in the SPA study, to test whether TBE rates correlate with atypical tactile responsivity. Again here, we focused on tactile responsivity given previously demonstrated associations between TBE and tactile responsivity^22, 24^. We evaluated the association between atypical tactile processing (overall Touch, Tactile Hyper- and Hypo-responsivity, Table S4) and TBE rates for the ASD (n=22) and TD (n=20) groups separately to ensure our findings reflected sensory responsivity rather than group differences (since ASD is known to be associated with atypical sensory responsivity; Figure 2a). The overall Touch subscale and TBE rates were positively correlated in ASD (rho=.475, p=.026), but not TD (rho=−.36, p=.119). Tactile Hyper-responsivity correlated with TBE rates in the ASD group (rho=.559, adjusted p = .028; Figure 2b). There was no significant correlation between Tactile Hypo-responsivity and TBE rates for either group (Figure 2c). These findings demonstrate the relevance of TBE rate to a tactile Hyper-responsivity phenotype.

**Figure 2.**
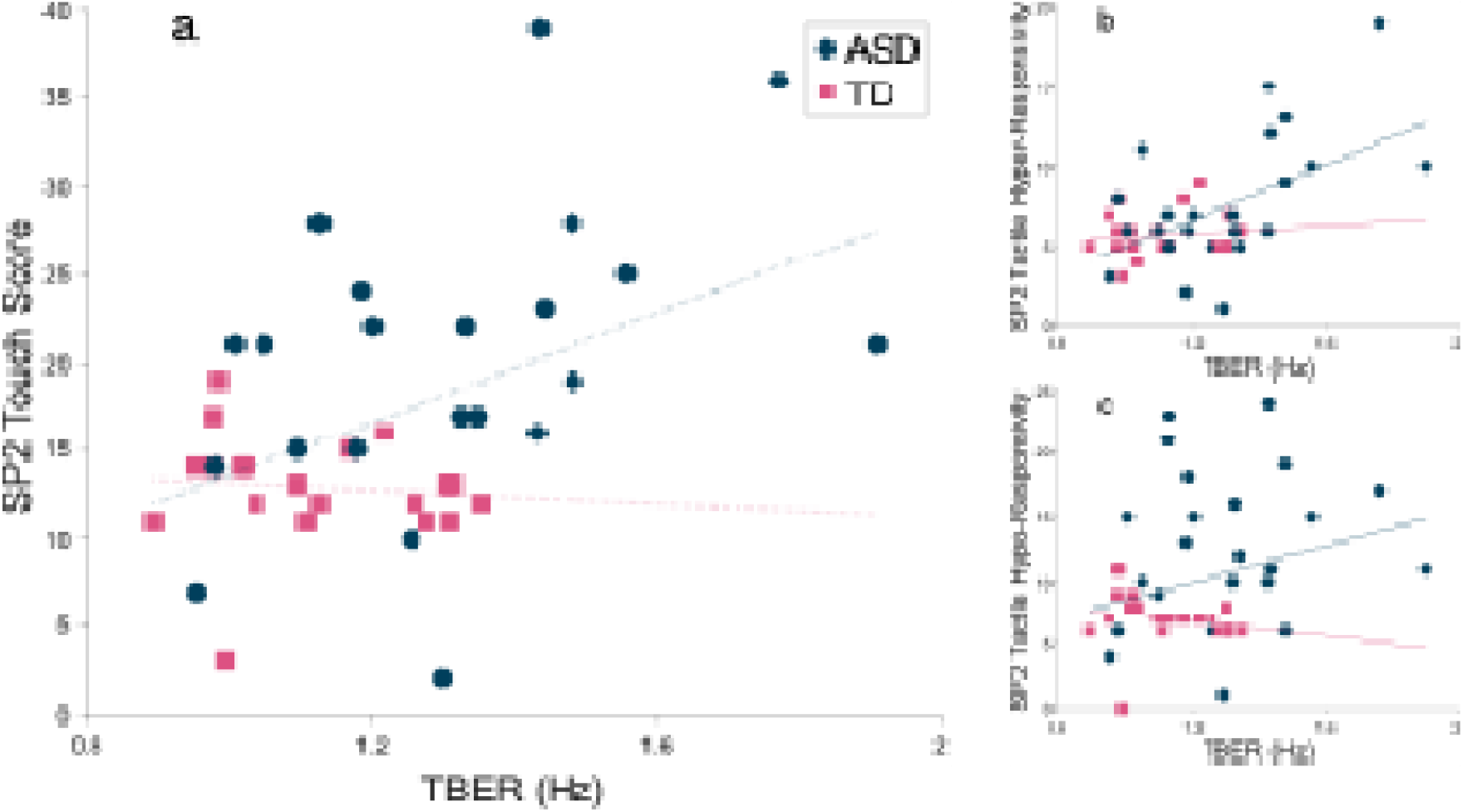
Association between TBE rate and SP2 tactile scores in SPA_ASD and SPA_TD cohorts. Each dot represents one participant. ASD blue circles; TD pink squares. Dashed lines reflect lines of linear best fit. r = Spearman’s Rank correlation coefficients. (a) TBE rates and SP2 Touch subscale were positively correlated in ASD (r = .475, p = .026), but not TD (r = −.360, p = 0.119). (b) TBE rates and Tactile Hyper-responsivity were positively correlated in ASD (r = .559, adjusted p = .028) but not TD (r = −.014, adjusted p = 1.000). (c) TBE rates and Tactile Hypo-responsivity were not significantly correlated for either group (ASD: r = .275, adjusted p = .859; TD: r = −.465, adjusted p = .156). (b,c) p-values Bonferroni corrected adjusting for 4 comparisons.

Neither age (rho = −.038, p = .360) nor Nonverbal Intelligence Quotient (NVIQ, rho = −.021, p=.611) were significantly associated with TBE rates. Medication likely does not account for high TBE rates as there was no consistent pattern of medications for participants with extreme TBE rates values (Table S5). TBE rates were not significantly higher based on seizure history (p=.150, Table S2).

Taken together, our finding that TBE rates are elevated in multiple rare neurogenetic conditions associated with atypical neurodevelopment (i.e. PMS, RTT, and *SYNGAP1*), and in a subset of children with idiopathic ASD, may reflect common circuit atypicalities and therapeutic targets across these conditions. Previous work identifying the thalamacortical mechanisms underlying TBEs ^21, 22, 24^, combined with our finding of elevated TBE rates in neurodiverse populations, suggests multiple future directions across the bench-to-bedside spectrum. For example, while thalamic bursting is sufficient to drive TBE ^22^, determining whether TBEs originate primarily in thalamus or are driven by more central (“top-down”) or peripheral (“bottom-up”) activity is important to identify targets for potential interventions and to develop TBE rates as a scalable, clinically useful stratifying biomarker and/or tool to monitor trial outcomes. Back-translation into animal models may be particularly useful for this purpose. Notably, shared treatments for tactile hyper-responsivity in mouse models of PMS and RTT are already being developed ^10^.

Our findings, specifically group differences between ASD patients relative to TD that likely reflect over-enrollment of children with sensory processing challenges in the SPA study, highlight the need to routinely characterize sensory processing challenges in studies of neurodevelopmental and neurogenetic conditions. While detailed sensory phenotyping of conditions as included here has begun in several studies ^36, 37^, many of the large consortia developing and testing biomarkers for these conditions did not initially include targeted sensory processing measures; including these in future study iterations, with careful consideration of recruitment biases, will be beneficial. While we focused on TBE over somatosensory cortex and its relationship to tactile processing given prior work, TBE are present across the scalp and more detailed future evaluations to localize these events, and examine their relationship to other sensory modalities and stimulus paradigms, will be important. Establishing baseline population TBE rate distributions will also be of useful to better understand why short-term reliability of TBE rates was higher in our ASD group. We suspect this is the result of low inter-individual variability, rather than high intra-individual variability, in the TD group, since ICC depends on both values.

Prior TBE studies focusing on individual trial level cortical responses to threshold-level stimuli demonstrated that the exact timing of stimulation relative to TBEs can create both hyper- and hypo-responsivity depending on the exact temporal relationship between TBEs and tactile stimuli^22^. Intrinsic brain activity (e.g., overall brain state, attention, TBE timing), sensory stimulus parameters (strength relative to detection threshold, salience, etc.), temporal context^38–40^, and perhaps spatial context can all contribute to dynamic, moment-to-moment fluctuations in sensory responsivity within individuals. Since TBEs are generated through the interplay of brief (∼50 ms) excitatory currents that recruit slower (100-300 ms) supragranular inhibition, it is reasonable to assume that total inhibition would increase through temporal integration relatively more than excitation as event rates increase. It is thus perhaps surprising then that we see a positive correlation between TBE rate and tactile hyperresponsivity rather than hyporesponsivity. However, is important to note that we measure tactile responsivity based on parent reports of naturalistic behavior. Future work designed to capture objective behavioral measures of sensory responsivity as a dynamic construct that varies from moment-to-moment within individuals, rather than characterizing hyper- vs. hyporesponsivity as a static clinical construct, will help address this issue.

In summary, TBEs reflect neural circuit activity with convergent and divergent properties across genetically-defined and phenotypically-defined (autism) disorders, respectively, offering potential as a mechanistically-based stratification biomarker. The extent to which TBEs reflect an aspect of daily function (tactile responsivity, with clear implications for quality of life in neurodivergent individuals) requires further investigation. With a known relation to underlying circuit dynamics, the TBE rate metric may inform better understanding, measurement, and eventual treatment of individuals in clinically and biologically meaningful subgroups of neurodevelopmental and neurogenetic conditions.

## Supplementary Figures and Tables

**Supplementary Table 1.**
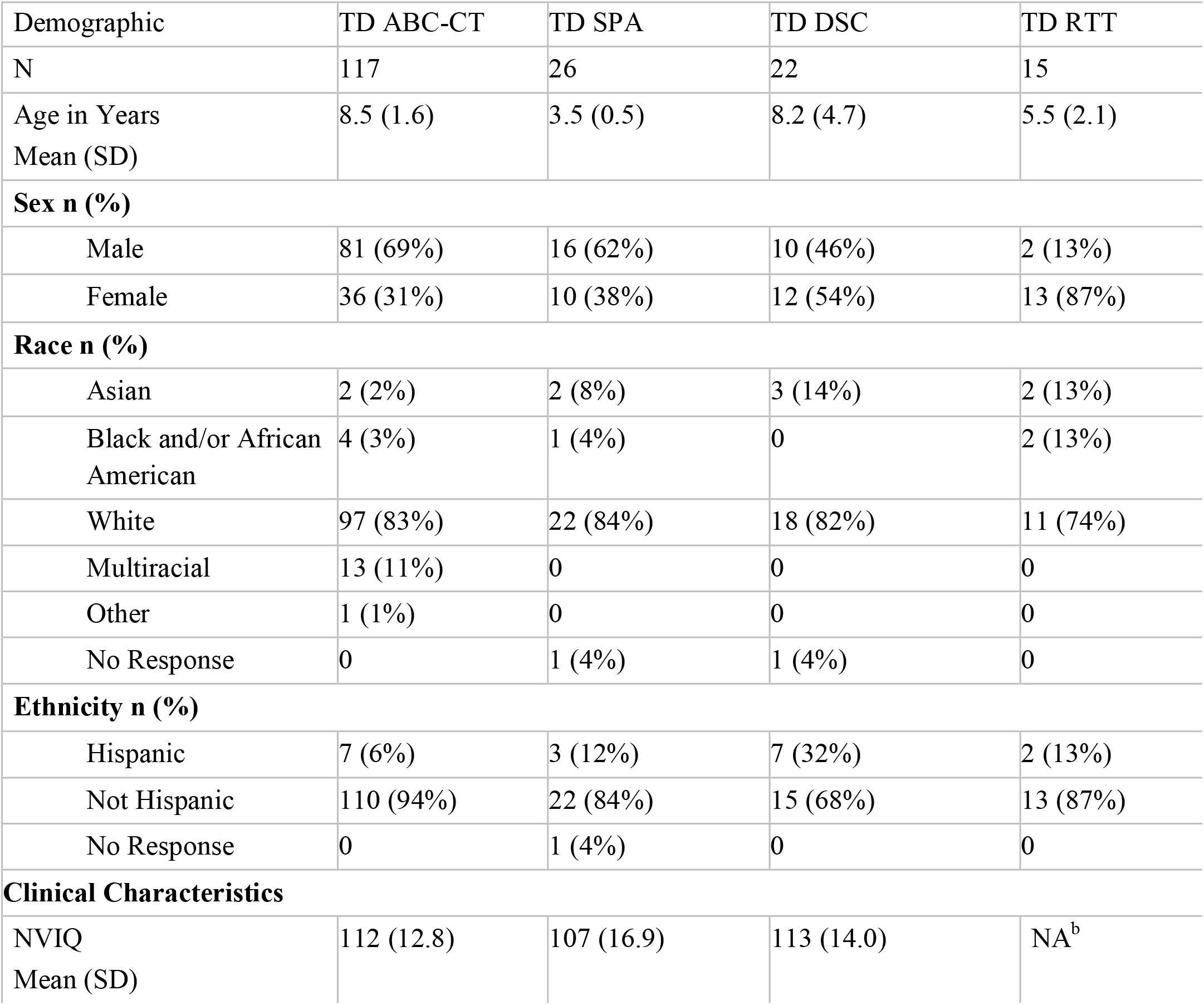
Demographics and Clinical Characteristics for TD groups from different studies.

| Demographic | TD ABC-CT | TD SPA | TD DSC | TD RTT |
| --- | --- | --- | --- | --- |
| N | 117 | 26 | 22 | 15 |
| Age in Years<br>Mean (SD) | 8.5 (1.6) | 3.5 (0.5) | 8.2 (4.7) | 5.5 (2.1) |
| **Sex n (%)** |  |  |  |  |
| Male | 81 (69%) | 16 (62%) | 10 (46%) | 2 (13%) |
| Female | 36 (31%) | 10 (38%) | 12 (54%) | 13 (87%) |
| **Race n (%)** |  |  |  |  |
| Asian | 2 (2%) | 2 (8%) | 3 (14%) | 2 (13%) |
| Black and/or African<br>American | 4 (3%) | 1 (4%) | 0 | 2 (13%) |
| White | 97 (83%) | 22 (84%) | 18 (82%) | 11 (74%) |
| Multiracial | 13 (11%) | 0 | 0 | 0 |
| Other | 1 (1%) | 0 | 0 | 0 |
| No Response | 0 | 1 (4%) | 1 (4%) | 0 |
| **Ethnicity n (%)** |  |  |  |  |
| Hispanic | 7 (6%) | 3 (12%) | 7 (32%) | 2 (13%) |
| Not Hispanic | 110 (94%) | 22 (84%) | 15 (68%) | 13 (87%) |
| No Response | 0 | 1 (4%) | 0 | 0 |
| **Clinical Characteristics** |  |  |  |  |
| NVIQ<br>Mean (SD) | 112 (12.8) | 107 (16.9) | 113 (14.0) | NA<sup>b</sup> |

**Supplementary Table 2.** Demographics and Clinical Characteristics for ASD, Neurogenetic, and Combined TD Groups.

|  | ASD<br>ABC-CT | ASD<br>SPA | PMS | TSC | PHTS | RTT | SYNGAP1 | TD |
| --- | --- | --- | --- | --- | --- | --- | --- | --- |
| N | 262 | 26 | 41 | 25 | 12 | 15 | 9 | 180 |
| Age in Years<br>Mean (SD) | 8.6 (1.6) | 3.7 (0.6) | 7.2 (3.5) | 6.7 (2.3) | 6.1 (2.6) | 8.4 (6.7) | 7.2 (2.3) | 7.5 |
| **Sex n (%)** |  |  |  |  |  |  |  |  |
| Male | 201 (77%) | 21 (81%) | 18 (44%) | 13 (52%) | 4 (33%) | 2 (13%) | 4 (44%) | 71 |
| Female | 61 (23%) | 5 (19%) | 23 (56%) | 12 (48%) | 8 (67%) | 13 (87%) | 5 (56%) | 109 |
| **Race n (%)** |  |  |  |  |  |  |  |  |
| Asian | 14 (5%) | 6 (23%) | 2 (5%) | 2 (8%) | 0 | 0 | 0 | 9 (5%) |
| Black and/or<br>African<br>American | 22 (8%) | 2 (8%) | 2 (5%) | 0 | 0 | 0 | 0 | 7 (4%) |
| White | 176 (67%) | 13 (50%) | 35 (85%) | 20 (80%) | 11 (92%) | 14 (93%) | 7 (78%) | 148 |
| Multiracial | 42 (16%) | 3 (11%) | 0 | 0 | 0 | 0 | 0 | 13 |
| Other | 6 (2%) | 0 | 2 (5%) | 0 | 0 | 1 (7%) | 0 | 1 (1%) |
| No Response | 2 (<1%) | 2 (8%) | 0 | 3 (12%) | 1 (8%) | 0 | 2 (22%) | 2 (1%) |
| **Ethnicity n (%)** |  |  |  |  |  |  |  |  |
| Hispanic | 47 (18%) | 7 (27%) | 3 (7%) | 5 (20%) | 2 (17%) | 2 (13%) | 1 (11%) | 19 |
| Not Hispanic | 215 (82%) | 17 (65%) | 38 (93%) | 19 (76%) | 10 (83%) | 13 (87%) | 6 (67%) | 160 |
| No Response | 0 | 2 (8%) | 0 | 1 (4%) | 0 | 0 | 2 (22%) | 1 (1%) |
| **Clinical Characteristics** |  |  |  |  |  |  |  |  |
| ASD diagnosis<sup>a</sup> | 262<br>(100%) | 26 (100%) | 25<br>(60.9%) | 7 (30.4%) | 5 (62.5%) | NA<sup>b</sup> | 7 (77.7%) | 0 |
| NVIQ<br>Mean (SD) | 98 (16.6) | 75 (24.8) | 44 (20.7) | 62.5 (34.9) | 72 (23.6) | NA<sup>b</sup> | NA<sup>b</sup> | 111 |
| Seizure History<br>n (%) | 0 | 0 | 7 (17%) | 21 (84%) | 1 (8%) | 12 (80%) | 8 (89%) | 0 |

<sup>a</sup> Percent ASD = ASD/ASD + No ASD. Missing data PMS n=0; TSC n=2; PHTS n=4; SYNGAP1 n=0.
<sup>b</sup> Not Available: No formal assessments were done in this domain.

**Supplementary Table 3.** TBE rates by Group.

| Diagnostic Group | TBE rates Median (IQR) |
| --- | --- |
| TD (N=180) | 1.10 (1.03 - 1.17) |
| ASD_SPA (N=26) | 1.26 (1.12 - 1.44) |
| ASD_ABC-CT (N=262) | 1.11 (1.05 - 1.18) |
| PMS (N=41) | 1.21 (1.12 - 1.59) |
| TSC (N=25) | 1.09 (0.86 - 1.44) |
| PHTS (N=12) | 1.07 (0.98 - 1.14) |
| RTT (N=15) | 1.52 (1.29 - 1.52) |
| *SYNGAP1* (N=9) | 1.40 (1.20 - 1.55) |

**Supplementary Table 4.** SP2 Tactile Measures.

| Diagnostic Group | Touch Domain Median (IQR) | Tactile Hyper-responsivity Median (IQR) | Tactile Hypo-responsivity Median (IQR) |
| --- | --- | --- | --- |
| TD (N = 20) | 13.0 (11-14) | 5.5 (5-6) | 7.0 (6-8) |
| ASD (N = 22) | 21.0 (15-24.5) | 6.5 (5-10.25) | 11.5 (8.25-16.25) |

**Supplementary Table 5.**
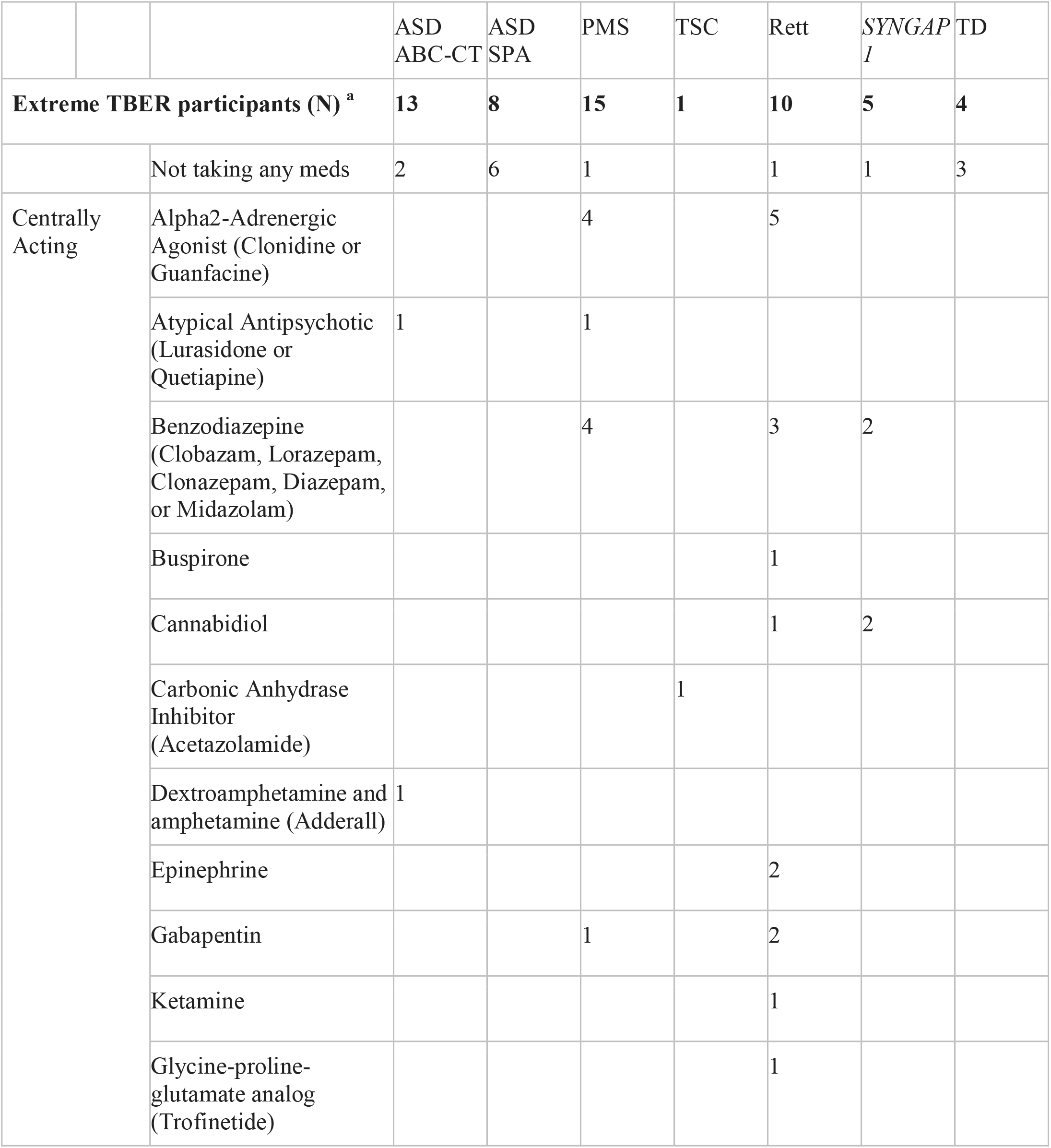
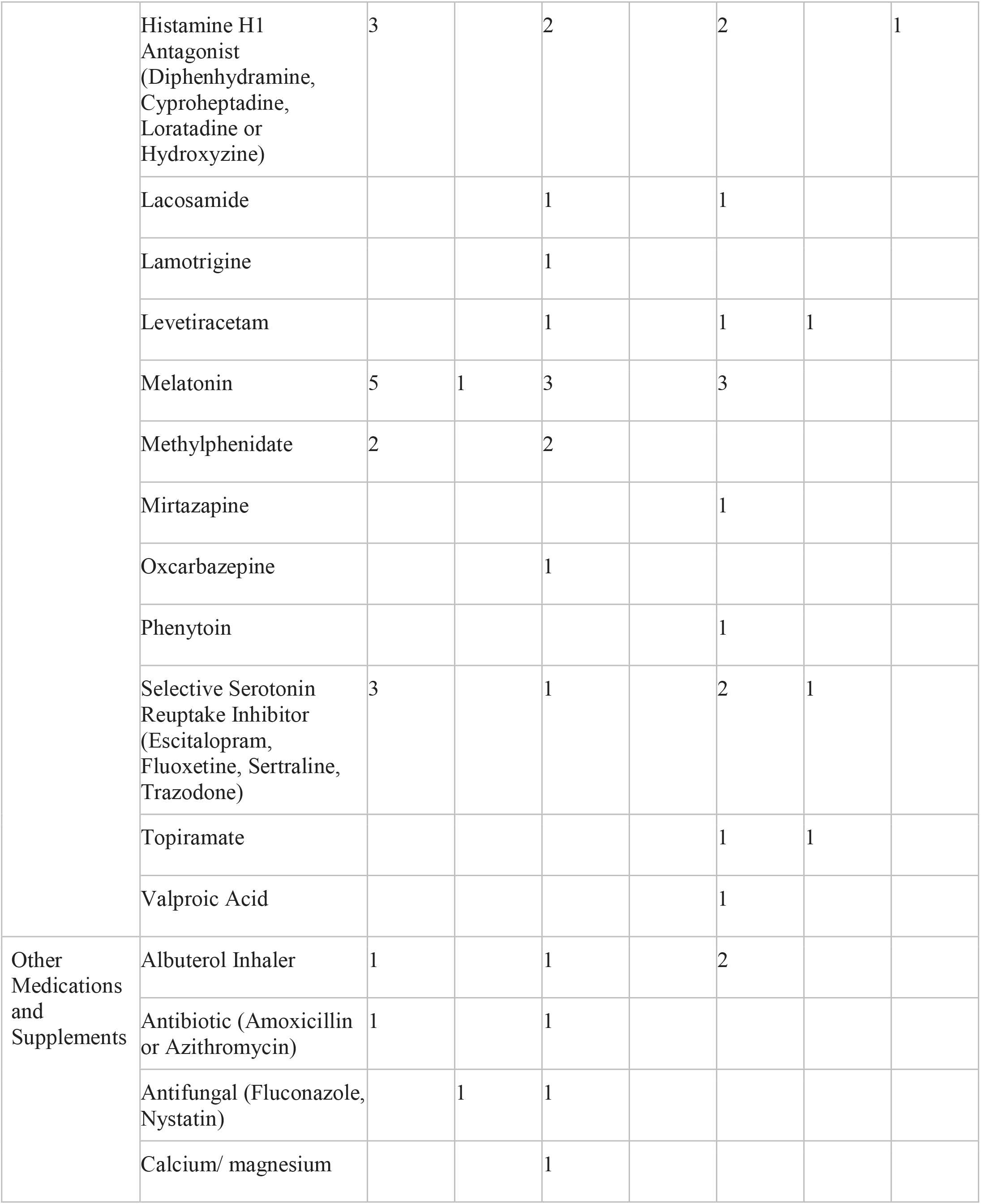
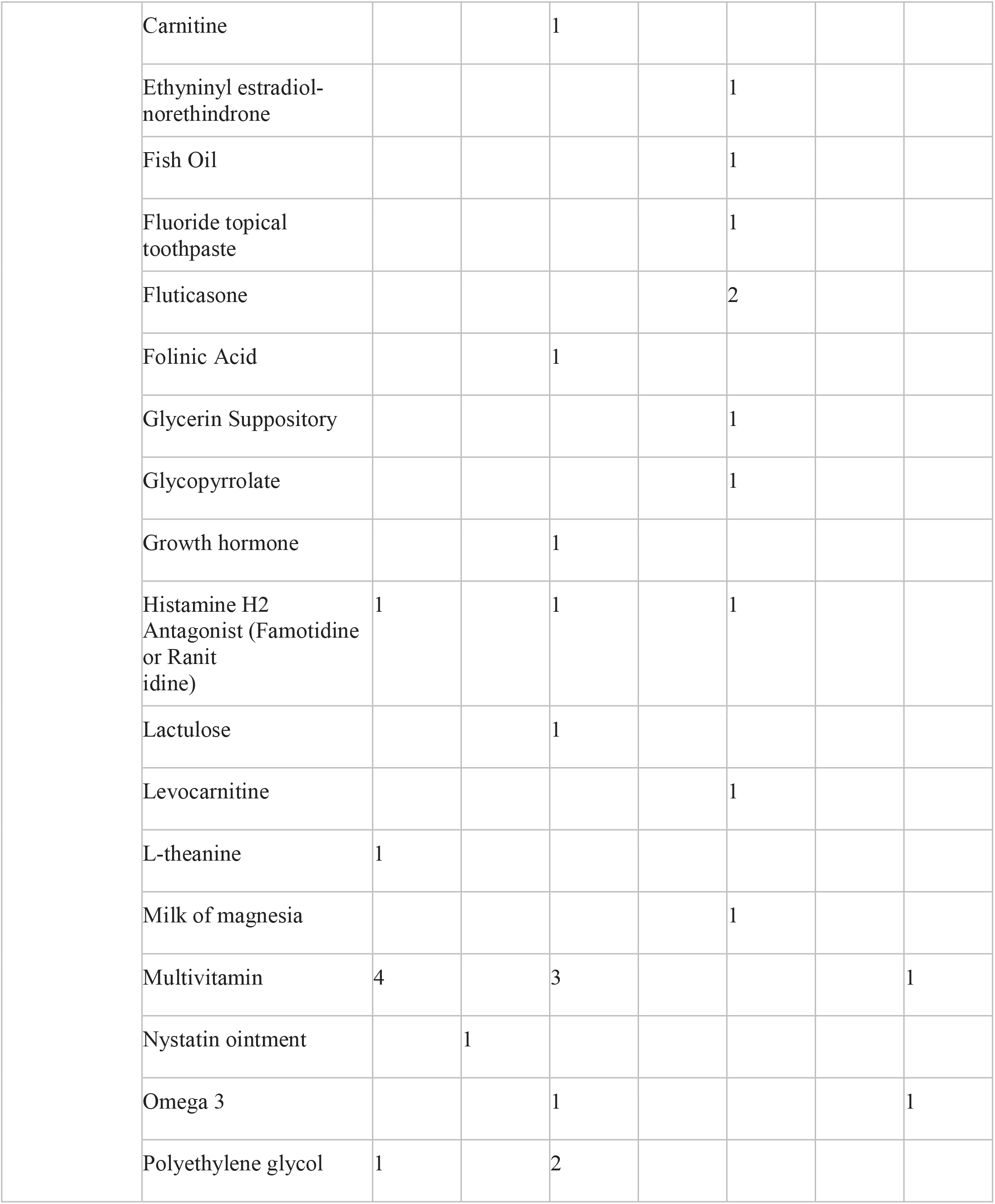
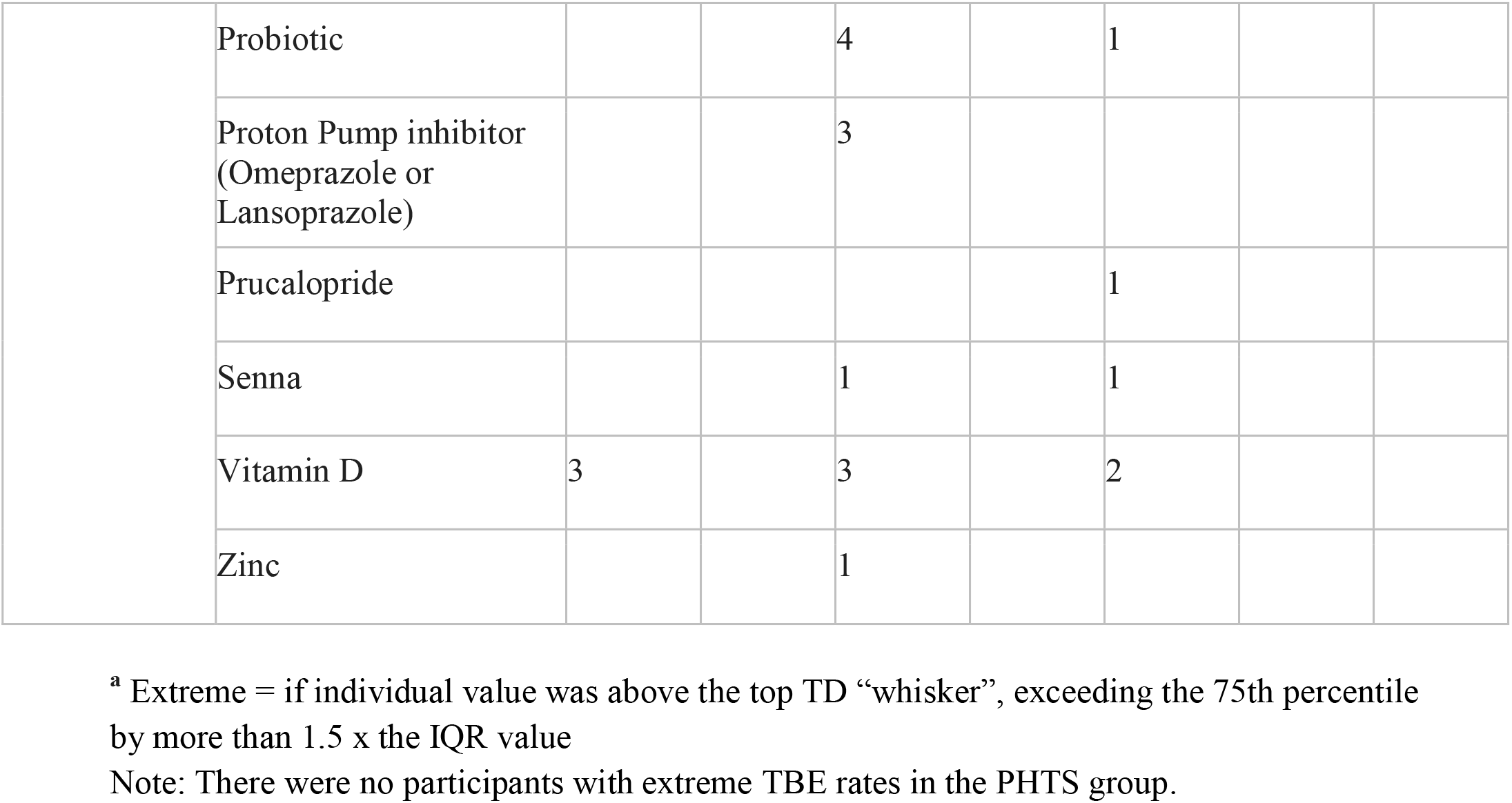
Medication List for participants with extreme TBE rates.

**Supplementary Table 6.**
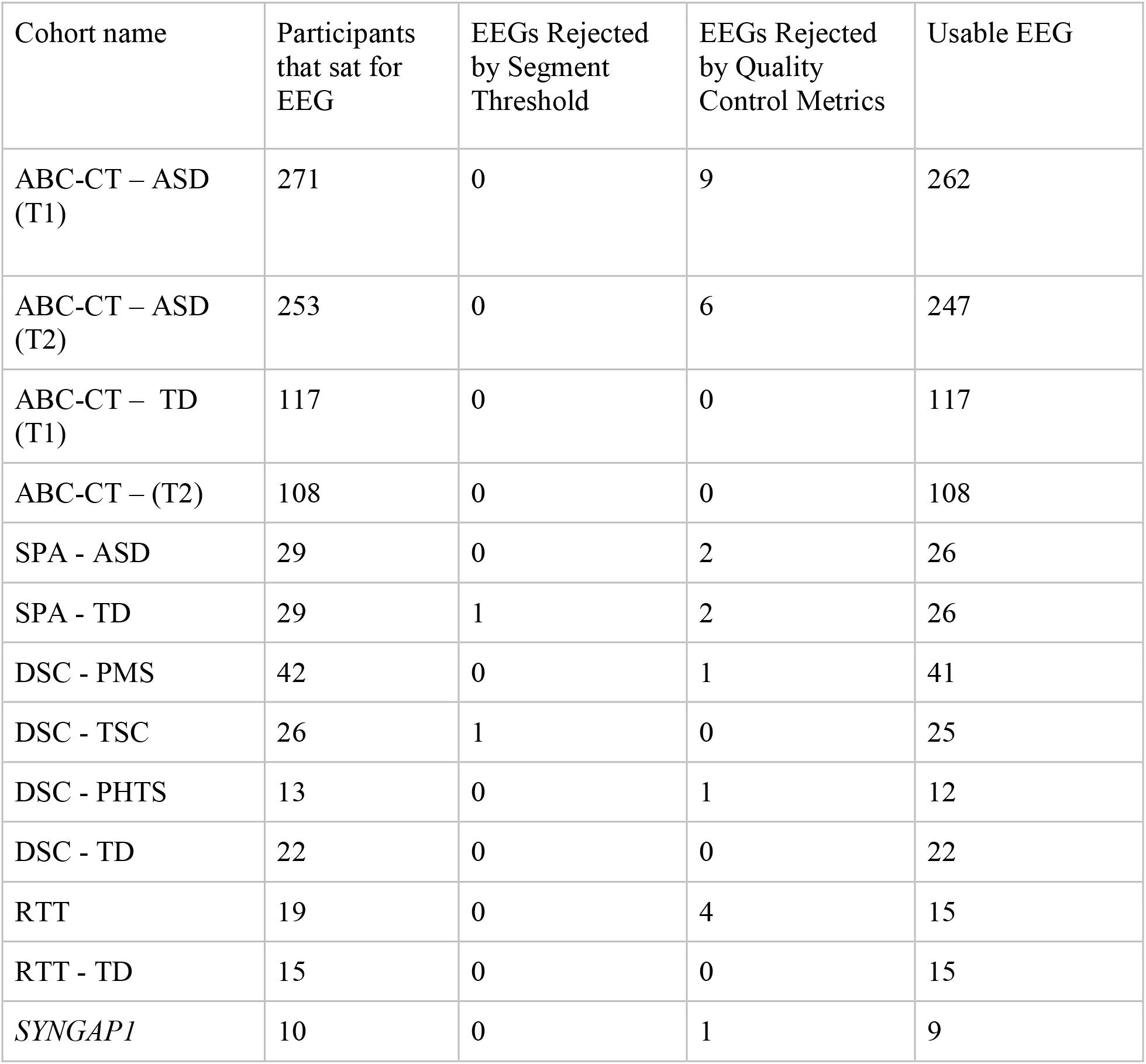
EEG Exclusion Breakdown by Cohort.

| Cohort name | Participants that sat for EEG | EEGs Rejected by Segment Threshold | EEGs Rejected by Quality Control Metrics | Usable EEG |
| --- | --- | --- | --- | --- |
| ABC-CT – ASD (T1) | 271 | 0 | 9 | 262 |
| ABC-CT – ASD (T2) | 253 | 0 | 6 | 247 |
| ABC-CT – TD (T1) | 117 | 0 | 0 | 117 |
| ABC-CT – (T2) | 108 | 0 | 0 | 108 |
| SPA - ASD | 29 | 0 | 2 | 26 |
| SPA - TD | 29 | 1 | 2 | 26 |
| DSC - PMS | 42 | 0 | 1 | 41 |
| DSC - TSC | 26 | 1 | 0 | 25 |
| DSC - PHTS | 13 | 0 | 1 | 12 |
| DSC - TD | 22 | 0 | 0 | 22 |
| RTT | 19 | 0 | 4 | 15 |
| RTT - TD | 15 | 0 | 0 | 15 |
| *SYNGAP1* | 10 | 0 | 1 | 9 |

**Supplementary Figure 1:**
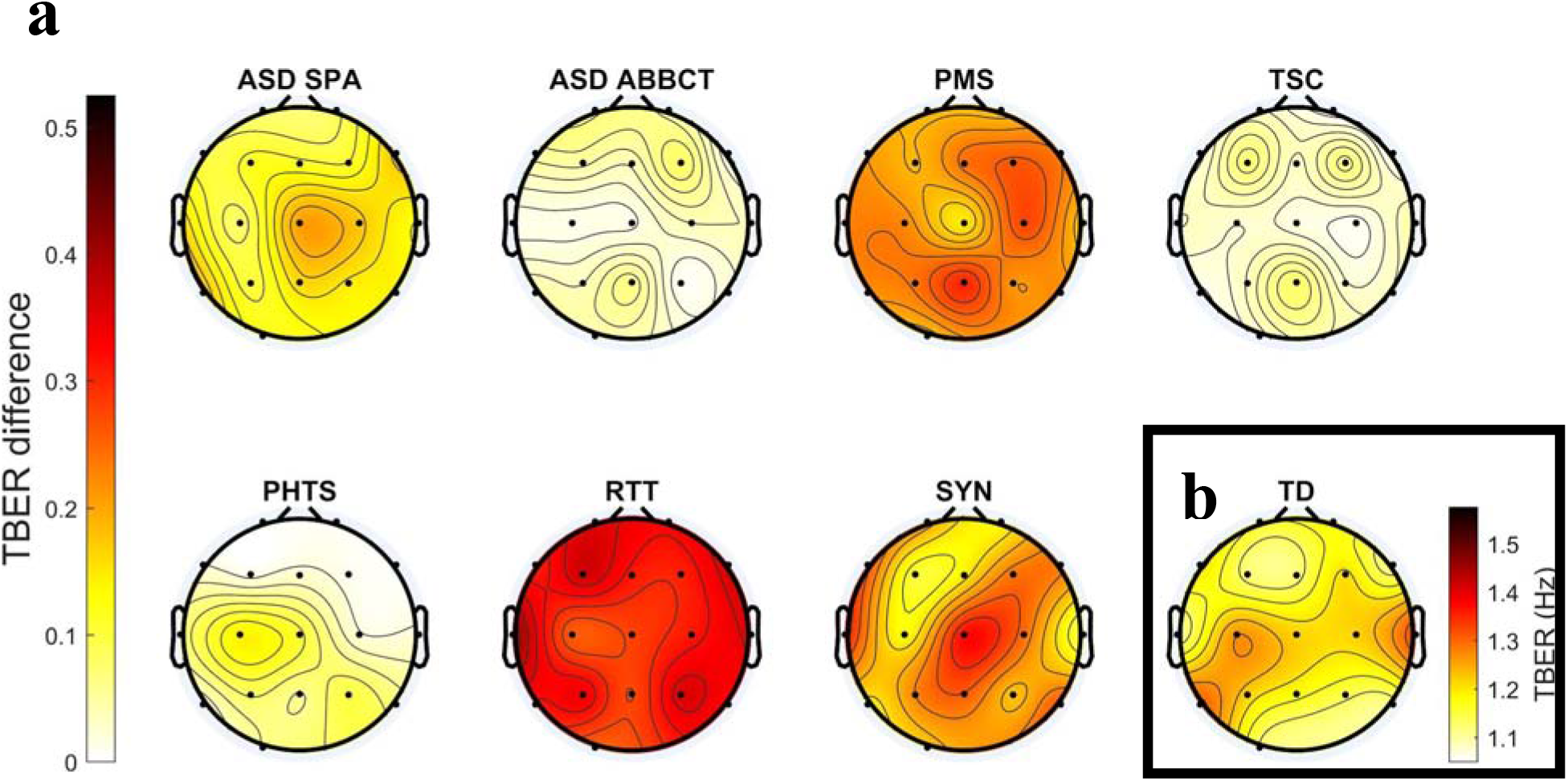
Topoplots showing (a) Difference in TBE rates (TBER, Hz) across the scalp (Cohort minus TD) and (b) TD TBER values.

**Supplementary Figure 2.**
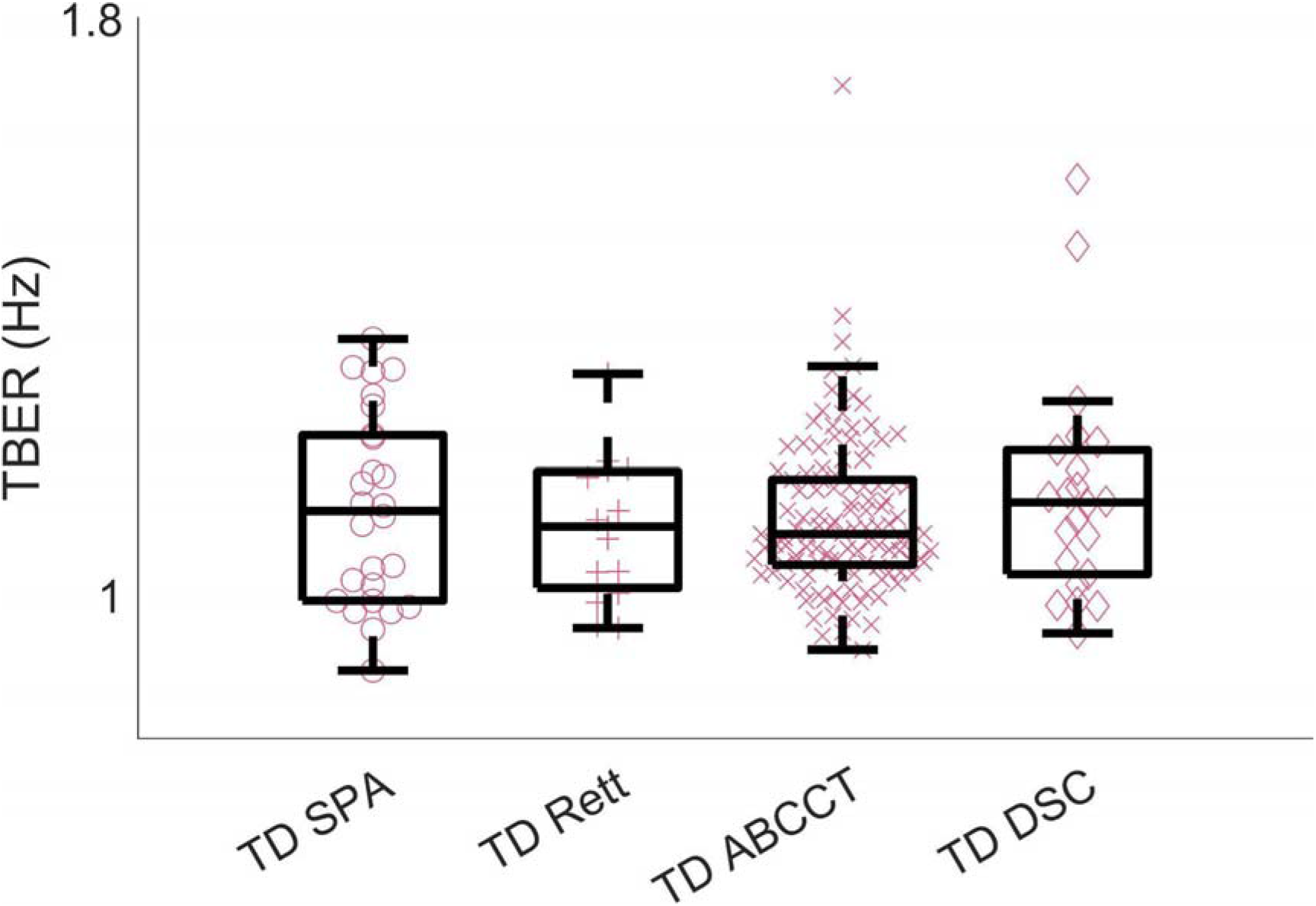
TD TBE rates Across Groups. Kruskal-Wallis test demonstrated no significant differences in TD groups across studies (p = .669).

**Supplementary Figure 3.**
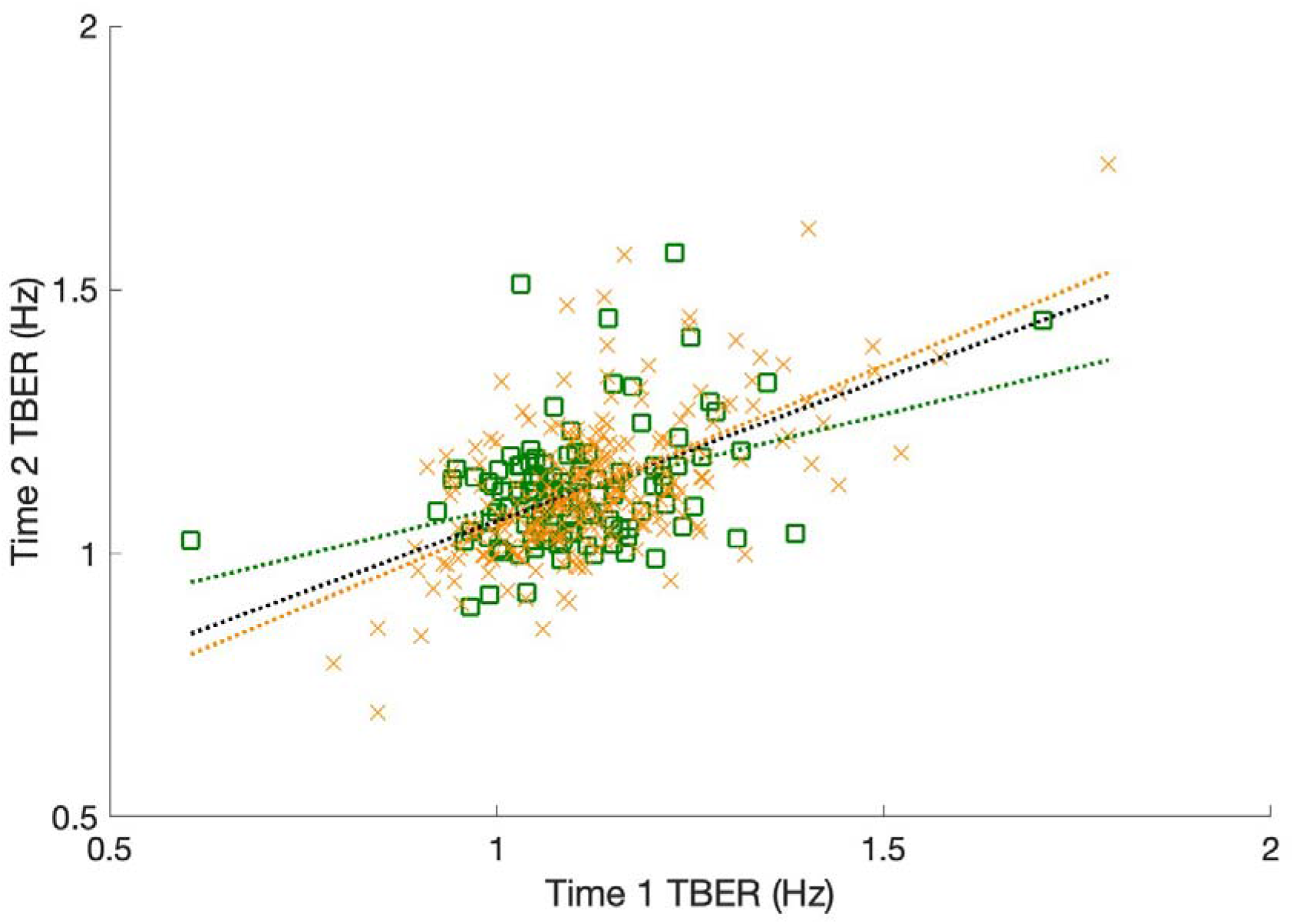
Short-term Reliability with ICCs for TD ABCCT (ICC = .365, 95% CI [.189, .517]) and ASD ABCCT (ICC = .606, 95% CI [.520, .679]) individually and collapsed into one group (ICC = .547, 95% CI [.470 .616]). Time 2 data was collected 6 weeks after Time 1. TD is green square, n =108; ASD is orange X, n = 247.

## Methods (Online Methods Section)

### Participants

Study participants were drawn from two studies of idiopathic ASD (note, genetic testing was not conducted in these studies) and three additional studies comprising five neurogenetic conditions commonly associated with ASD. Data and ethnicity is per report from the parent or legal guardian of each participant, using classification terms provided by the research team. The definition of “usable” EEG data is below in the section on “EEG preprocessing”, and quantification of EEGs excluded are described in Table S6.

### Autism Biomarkers Consortium for Clinical Trials (ABC-CT)

Data were collected from six- to eleven-year-old TD and ASD children across two visits, six weeks apart, from five sites: Boston Children’s Hospital; University of California, Los Angeles; University of Washington; Duke University; and Yale University. ASD diagnoses were confirmed using rigorous standardized diagnostic measures (Autism Diagnostic Observation Schedule, Second Edition (ADOS-2 ^41^ and DSM-5 criteria by reliable clinicians). Study details have been previously described ^35, 42^. 117 TD and 262 ASD participants provided usable EEG data at the first timepoint (T1), and of these 108 TD and 247 ASD participants provided usable EEG data at the six-week timepoint (T2, see Table S6). Exclusionary criteria included neurological conditions (including epilepsy), known genetic conditions causally related to ASD, or full scale IQ that fell outside of 60-150 on the Differential Ability Scales, Second Edition (DAS-II ^43^). Medication was allowed given a stable regimen of at least eight weeks preceding enrollment. TD participants were also excluded if they had a sibling with ASD or if they had an active psychiatric disorder. The protocol was approved and overseen by a central IRB at Yale University (HIC#: 1509016477; FWA00002571). Written informed consent from all guardians, and assent for all participants, was obtained after procedures were fully explained and the opportunity to ask questions offered.

### Sensory Processing and Adaptation (SPA) Study

Data were collected from three- and four-year-old TD and ASD children at Boston Children’s Hospital at a single time point. 26 TD and 26 ASD participants provided usable EEG data. Group placement was confirmed by the ADOS-2 ^41^ conducted by a research reliable examiner, as well as structured developmental history. Mullen Scales of Early Learning ^44^ were conducted to confirm that social communication challenges could not be better accounted for by cognitive or language delays. Exclusionary criteria included deafness, blindness, inability to complete procedures in English, nonverbal age less than 18 months on the Mullen, and motor challenges that preclude participation in testing. TD participants were excluded if they had neurodevelopmental concerns or a first-degree relative with ASD. The protocol was approved and overseen by the IRB at Boston Children’s Hospital (IRB-P00029639). Informed consent from all guardians, and assent for all participants developmentally able to assent, was obtained after procedures were fully explained and the opportunity to ask questions offered.

### Developmental Synaptopathies Consortium (DSC)

Data were collected from two to 11 year old children with TD, Phelan McDermid Syndrome (PMS), Tuberous Sclerosis Complex (TSC), and *PTEN* Hamartoma Tumor Syndrome (PHTS) at one time point from seven sites (Boston Children’s Hospital; University of California, Los Angeles; Cincinnati Children’s Medical Hospital, Cleveland Clinic, National Institute of Health and Icahn School of Medicine at Mount Sinai). 22 TD, 25 TSC, 41 PMS and 12 PHTS provided usable EEG data. Exclusionary criteria included taking an investigational drug as part of another research study within 30 days prior to study enrollment, behaviors that could prevent EEG lead placement and data collection, and inability to comply with study procedure/assessments. Additional exclusions for typically developing controls were uncorrected auditory or visual impairments. Autism diagnostic confirmation was provided by Clinician Best Estimate informed by gold-standard measures including the Autism Diagnostic Interview-Revised (ADI-R)^45^, ADOS-2 ^41^ (Childhood Autism Rating Scale-2, CARS-2 ^46^, for telehealth visits during the COVID-19 pandemic), DSM-5 Checklist ^5^, medical history, and clinical certainty ratings. In cases where data was unavailable or invalid (e.g., due to extremely low cognition), information on autism diagnosis was marked as “missing.” The protocols were approved and overseen by a central IRB at Boston Children’s Hospital (PMS HIC#: IRB-P0001330; TSC HIC#: IRB-P00013585; PHTS HIC#: IRB-P00013150). Informed consent from all guardians, and assent for all participants where cognitively and developmentally feasible, was obtained after procedures were fully explained and the opportunity to ask questions offered.

### SYNGAP1-Related Disorder (SYNGAP1)

Data were collected from two to 11 year old *SYNGAP1* participants at one timepoint from two sites: Boston Children’s Hospital and University of California, Los Angeles. Nine *SYNGAP1* participants provided usable EEG data. Exclusion criteria centered on whether the *SYNGAP1* variant could be validated by the Ciitizen *SYNGAP1* Natural History registry (https://www.ciitizen.com/syngap1). Autism diagnosis was based on parent report of whether the child had previously received an autism diagnosis from a clinician. The protocol was approved and overseen by a central IRB at Boston Children’s Hospital (IRB-P00039324). Informed consent from all guardians, and assent for all participants developmentally able to assent, was obtained after procedures were fully explained and the opportunity to ask questions offered.

### *Rett* Syndrome

Data were collected from one to 25 year old TD and RTT participants at one time point at Boston Children’s Hospital. 15 TD and 15 RTT participants provided usable data. In the RTT group, inclusion criteria included a diagnosis of RTT with a previously identified pathogenic or likely pathogenic variant in the methyl-CpG binding protein 2 (*MECP2)* gene. Exclusionary criteria included taking an investigational drug as part of another research study within 30 days prior to study enrollment. Additional exclusion criteria for the TD group included no diagnoses or medical conditions associated with increased risk of ASD or intellectual disability. ASD diagnostic status was not included in this study as limited motor skills in many of the participants precluded accurate assessment of developmental and cognitive level which in turn precluded assessment of whether social communication skills were impaired out of proportion to cognitive skills, as would be necessary to determine ASD diagnostic status. The protocol was approved and overseen by the IRB at Boston Children’s Hospital (IRB-P00038294). Informed consent from all guardians, and assent for all participants developmentally able to assent, was obtained after procedures were fully explained and the opportunity to ask questions offered.

Demographics and clinical characteristics for all cohorts can be found in Table S1 and S2.

### Phenotypic Data: Sensory Profile 2

SPA participants completed the Sensory Profile 2 Caregiver questionnaire (SP2) ^47^ a norm-referenced questionnaire that evaluates a child’s sensory processing across home, school, and community contexts (SPA_ASD n = 22; SPA_TD n = 20; Table S4). For the current study, three summary scores were extracted: the overall Touch Subscale, and Tactile Hyper-Responsivity and Tactile Hyper-Responsivity scores. As previously described ^8^, the Tactile Hyper-Responsivity score was the sum the following items (each scored on a Likert scale from 1 to 5): “Shows distress during grooming,” “Becomes irritated by wearing shoes or socks,” “Shows an emotional or aggressive response to being touched,” “Becomes anxious when standing close to others,” and “Rubs or scratches a part of the body that has been touched.” The tactile Hypo-Responsivity score was the sum of the following items: “Touches people or objects to the point of annoying others,” “Displays need to touch toys, surfaces, or textures,” “Seems unaware of pain,” “Seems unaware of temperature changes,” “Touches people and objects more than same-aged children,” and “Seems oblivious to messy hands or face.” Of note, some studies have called this “Hyper-Reactivity” ^8^ but we use the term “Hyper-Responsivity” to better reflect behavioral responsivity as recommended by He et al ^48^.

### Nonverbal Intelligence Quotient (NVIQ)

To assess non-verbal cognitive ability, participants in the ABC-CT, SPA, and DSC cohorts were administered either the Mullen Scales of Early Learning ^44^, the Differential Ability Scales, 2nd edition (DAS-II) ^43^, or the Stanford Binet-5 (SB-5) ^49^. NVIQ was compiled based on the test given: for the Mullen Scales of Early Learning, this was derived from fine motor and visual reception developmental quotients. For the SB-5, the NVIQ was taken directly. For participants given the DAS-II, the nonverbal reasoning standard score was taken. No formal cognitive assessments were done as part of the RTT study (due to concerns that motor abilities may preclude accurate assessment) or *SYNGAP1* study.

### EEG Data Collection

EEG data were collected using 128 or 64 channel montages (see below) in the resting condition using similar protocols across all studies, as described below. Given the variable abilities of study participants to follow instructions, “resting” EEG is defined here as data collected during an awake quiet state with eyes open and without any time-locked stimulus or task, consistent with standards in the autism field^42^.

### ABC-CT

EEG data were collected in full lighting, using a 128-channel Hydrocel Geodesic Net (Electrical Geodesic, Inc., Eugene, OR) and NetStation software (Geodesic, Inc) with a sampling rate of 1000 Hz. Children watched an approximately three-minute video, comprising six soundless videos akin to screensavers. Each video played forward for 15 seconds and then in reverse for 15 seconds. The six videos were played in three blocks of two videos each at 30 frames per second. Video display was limited to 7 × 9.3 cm to minimize eye movement during task completion. A behavioral assistant sat in the room with the participant to direct the child to pay attention to the screen.

### SPA

EEG data were collected in dim lighting, and methods were otherwise identical to those described for the ABC-CT cohort.

### DSC

EEG data were collected in dim lighting, and for the Cleveland Clinic site, was recorded on Brain Vision system with Brain Vision 64 channel cap. Methods were otherwise identical to ABC-CT.

### RTT

Children watched an extended version of the video described above, lasting for approximately 5 minutes. Methods were otherwise identical to the SPA and *SYNGAP1* cohorts.

### SYNGAP1

EEG data were collected in dim lighting, and methods were otherwise identical to those described for the ABC-CT cohort.

### EEG Pre-Processing

EEG data from all cohorts were exported to MATLAB and pre-processed using the Batch EEG Automated Processing Platform ^50^. Data were filtered using a bandpass filter from 0.1-100 Hz and resampled to 250 Hz. Artifact detection and correction was completed using the Harvard Automated Preprocessing Pipeline for EEG (HAPPE) pipeline, which is optimized for short recording EEG data collected from children with neurodevelopmental conditions ^51^. HAPPE applies line noise removal at 60 Hz, then detects and removes bad channels and artifacts in the data, such as eyeblinks and eye or muscle movements, using wavelet-enhanced independent component analysis (w-ICA) and ICA with the multiple artifact rejection algorithm (MARA ^52, 53^. The following channels, in addition to the 10–20 electrodes, were used for ICA with MARA: FC3, CP3, C1, C5, FC4, CP4, C2, C6, AF3, F1, AFz, AF4, F2. Electrodes were spread evenly across the scalp, and the number of electrodes was chosen relative to our recording length to maximize ICA performance and prevent overfitting of the algorithm ^50, 54^. Following artifact removal, bad channels were interpolated, and data were re-referenced to an average reference. Data were then segmented into 5□s windows. Segments were inspected again for artifact and segments rejected using HAPPE’s recommended amplitude cutoff (40 μV cutoff) and joint probability criteria. The 40□μV cutoff reflects the smaller amplitude that results from the wavelet thresholding and ICA steps during artifact detection^51^.

### EEG Rejection Criteria

EEGs were rejected if they had fewer than 25 segments (125 sec) or fell outside of 3 standard deviations from the mean across participants on the following HAPPE quality control parameters: percent good channels (3 SD: < 76%), mean retained artifact probability (3 SD: > 0.27), median retained artifact probability (3 SD: > 0.25), percent of independent components rejected as artifact (3 SD: > 75%), and percent of EEG signal variance retained after artifact removal (3 SD: < 15%). Based on the above criteria 20 of 594 (3.4%) EEGs collected were rejected.

For all participants who had more than 25 segments available, Spearman’s correlation revealed no significant association between segment number and TBE rates (p = .864). See Table S6 for a summary of EEG exclusion by cohort. “Usable” EEGs for each cohort are defined as EEGs that were collected and not rejected based on the above criteria.

### Transient Beta Event Detection and Event Rate Calculation

Transient beta events were detected in single trial segmented data from using the SpectralEvents Toolbox ^24^. Each 5-second segment was convolved with 5-cycle Morlet wavelets to calculate the time-frequency response (TFR) every 4 Hz between 13 and 30 Hz ^55^. Each TFR was then normalized by the median power in each frequency; thus, units of the normalized TFR are in factors of median. Transient events were then detected using the “minimal overlap” option of the toolbox (find method 2); local maxima were found in the normalized TFR using MATLAB’s imregionalmax function (R2021a). The TFR was thresholded to a cutoff of 6 factors of median ^24^, and the center of each beta event was then defined as the single local maximum in each suprathreshold region of the TFR.

The rate of event occurrence was found in each segment by counting the number of detected events and dividing by 5, in order to yield an event rate in Hertz for each 5-second segment. For each participant, the TBE rates was defined as the average of this value across all segments of a channel’s data. The averaged TBE rates in channels C3 and C4 for each individual were used for all analyses because prior studies of TBE have primarily focused on somatosensory cortex ^23, 24^. Topoplots (Figure S1) show TBE rates in other channels.

### Statistical Analyses

Prior to analyses, we confirmed that TBE rates did not vary for TD participants across cohorts using a Kruskal-Wallis test (Figure S2) and therefore collapsed all TD participants into one group. Because our EEG data were non-normally distributed, we used non-parametric tests for all EEG analyses. All analyses were performed by IBM SPSS Statistics version 27 (IBM Corp).

### Comparing Transient Beta Event Rate

Provided (1) we only had sensory phenotyping data for ASD participants in the SPA cohort and (2) study description in SPA may have self-selected for participants with elevated sensory processing difficulties, the ASD participants from the SPA and ABC-CT cohorts are treated as separate groups (hereafter referred to as ASD_SPA and ASD_ABC-CT). To compare transient beta event rate, we tested whether TBE rates at C3/C4 in ASD participants (ASD_SPA and ASD_ABC-CT) differed significantly from our collapsed TD group via two independent samples Mann-Whitney U tests.

We next evaluated TBE rates in a variety of neurogenetic conditions, all of which increase the likelihood of ASD. Specifically, we used five Mann-Whitney U tests, Bonferroni corrected for five comparisons, to evaluate whether TBE rates in TD participants significantly differed from TBE rates in each neurogenetic group (PMS, TSC, PHTS, RTT, *SYNGAP1*).

### Evaluate the short-term reliability of TBE rates across two time points

Since evaluation of TBE rates in EEG and in neurodevelopmental conditions more generally is a novel analysis, and with an eye towards potential utility of TBE rates as a clinically relevant biomarker of atypical sensory processing in these conditions, we also wished to assess the short-term reliability of TBE rates. For participants in the ABC-CT cohort with usable EEG data at both visits, we ran intraclass correlations (ICC) by diagnostic group to assess the short-term reliability of TBE rates. ICC values were interpreted as poor (< 0.4), fair (0.4 to .59), good (0.6 to 0.74), or excellent (0.75 to 1.0) ^56^.

### Evaluate the relationship between TBE rates and Tactile Sensory Processing

For participants who provided sensory data (from the SPA study), we evaluated the relationship between TBE rates and tactile processing using Spearman’s rank correlations for the ASD and TD groups separately. Our reason for examining groups separately is that ASD is known to be associated with atypical sensory responsiveness and we wanted to ensure our findings reflected sensory responsiveness rather than differences in diagnostic groups. First, we correlated TBE rates with the SP2 touch domain score. Next, to evaluate how TBE rates related to sub-domains of touch, we carried out four separate Spearman’s rank correlations to test the relationship between TBE rates and Tactile Hyper-responsivity and Tactile Hypo-responsivity, within the ASD and TD groups separately. For these subdomain analyses, we thus use p-values Bonferroni corrected for 4 comparisons.

### Evaluate the relationship between TBE rates and other demographic and clinical characteristics

We hypothesized a positive association between TBE rates and tactile responsivity, but also evaluated whether other demographic and clinical characteristics (age, NVIQ, epilepsy, medications, ASD) could account for our findings. We used Spearman’s rank correlations to evaluate the relationship between TBE rates and age for all participants. We used Spearman’s rank correlations to evaluate the relationship between TBE rates and NVIQ for all participants in whom NVIQ data were available (ABC-CT, SPA, DSC). Across all neurogenetic conditions that measured ASD (DSC, *SYNGAP1*), we used one Mann-Whitney U-test to evaluate whether TBE rates differed in participants with or without co-occurring ASD. Since neurogenetic conditions had higher rates of seizures than idiopathic ASD, across all neurogenetic conditions we compared TBE rates in participants with versus without history of seizures using a Mann-Whitney U-test. To determine whether medications could contribute to particularly elevated TBE rates, the small numbers of participants taking a wide variety of medications precluded direct statistical comparisons. Instead, we compiled medication lists for participants with “extreme” TBE rates. TBE rates were defined as “extreme” if it was an outlier based on the TD group’s threshold (i.e., if it fell above the top TD “whisker”, exceeding the 75th percentile by more than 1.5 x the IQR value). (Table S5).

## Data availability statement

The data that support the findings of this study are available, in collaboration with the principal investigator for each respective study, upon reasonable request. Data from the Autism Biomarkers Consortium for Clinical Trials are also available via the NIMH Data Archive (identifier: 2288; https://nda.nih.gov/edit_collection.html?id=2288).

## Code Availability statement

EEG preprocessing was conducted using the Batch EEG Preprocessing Platform (BEAPP), available at https://github.com/lcnbeapp/beapp. Within BEAPP we used the Harvard Automated Preprocessing Pipeline for EEG (HAPPE), which is embedded in BEAPP and also available separately at https://github.com/PINE-Lab/HAPPE. To calculate TBE rates we used the SpectralEvents Toolbox, available at https://github.com/jonescompneurolab/SpectralEvents.

## Acknowledgements

This work was supported by grants from the Simons Foundation/SFARI (Award number 648277, ARL), the Hock E. Tan and K. Lisa Yang Center for Autism Research at Harvard University (ARL), the Clark Family (ARL/MF), the Eagles Autism Foundation (ARL), the National Institute of Mental Health (U19 MH108206, JCM; R01 MH122428, DŞ), Intramural Research Program ZICMH002961, National Institute of Neurological Disorders and Stroke (R01 NS134948, ARL; U54 NS092090, MS), National Institute of Child Health and Human Development (P50HD105351, MS; P50HD105351 Project 001, MF), and CURE *SYNGAP1* (ARL/MS/AP/KW). We are grateful to all the children and families who participated in these studies.

The Developmental Synaptopathies Consortium acknowledges the support of Simon K. Warfield, Benoit Scherrer, Rajna Filip-Dhima, Kira Dies, Paige Siper, Ellen Hanson, and Jennifer M. Phillips.

The Autism Biomarkers Consortium for Clinical Trials acknowledges the support of Madeline Aubertine, Jessica Benton, Cynthia Brandt, Carter Carlos, Shou-An A. Chang, Kelsey Dommer, Alyssa Gateman, Simone Hasselmo, Julie Holub, Toni Howell, Ann Harris, Alexander Hoslet, Kathryn Hutchins, Kelsey Jackson, Scott Johnson, Lily Katsovitch, Minah Kim, Beibin Li, Kelsey MacDonald, Samantha Major, Samuel Marsan, Adriana S. Méndez Leal, Takumi McAllister, Lisa Nanamaker, Leon Rozenblit, Megha Santosh, Helen Seow, Laura Simone, Dylan Stahl, Cindy Voghell, Andrew Yuan, and Taylor Hoffman. Consultation for the ABC-CT EEG was provided by the EU Aims LEAP team, including Declan Murphy, Eva Loth, Emily J.H. Jones and Luke Mason.

